# Effects of sub-anesthetic oro-mucosal dexmedetomidine on sleep in humans: A pharmacokinetics-pharmacodynamics study

**DOI:** 10.1101/2024.07.03.24309892

**Authors:** Laura K. Schnider, Marta Ratajczak, Rafael Wespi, Jacqueline G. Kientsch, Francesco Bavato, Laurenz Marten, Jonas Kost, Maxim Puchkov, Corinne Eicher, Martina Boxier, Clarissa D. Voegel, Oliver G. Bosch, Eus van Someren, Dario A. Dornbierer, Hans-Peter Landolt

**Affiliations:** Institute of Pharmacology & Toxicology, University of Zurich, Zurich, Switzerland; Department of Adult Psychiatry and Psychotherapy, University Hospital of Psychiatry, University of Zurich, Zurich, Switzerland; Department of Pharmaceutical Sciences, University of Basel, Basel, Switzerland; Institute of Forensic Medicine, University of Zurich, Zurich, Switzerland; Center for Forensic Hair Analytics, Zurich Institute of Forensic Medicine, University of Zurich, Zurich, Switzerland; Netherlands Institute for Neuroscience, Amsterdam, The Netherlands; Sleep & Health Zurich, University of Zurich, Zurich, Switzerland

## Abstract

**Background:** The locus coeruleus noradrenergic (LC-NE) system may provide a potential new target for pharmacological insomnia treatment, particularly in patients suffering from elevated stress. The selective α_2_ noradrenergic agonist dexmedetomidine (DEX) attenuates LC-NE activity in sub­ anesthetic doses, yet no adequate non-parental delivery systems of DEX are currently available. To examine the feasibility of oro-mucosal DEX administration, we developed two distinct – one sublingual and one buccal – oro-mucosal, fast-disintegrating DEX formulas tailored for self­ administration. Here we established their pharmacokinetic and pharmacodynamic (PK-PD) profiles.

**Methods:** In two separate studies in 8 male good sleepers and 17 men with subclinical insomnia, we administered sub-anesthetic doses (20 & 40 µg) of the two formulas following a randomized, double-blind, placebo-controlled, cross-over design. We complemented the PK assessments with all­ night polysomnography, nocturnal cortisol and melatonin measurements, assessments of cardiovascular functions during and after sleep, cortisol awakening response, and post-awakening examination of subjective state and vigilance.

**Results:** Particularly buccal DEX was rapidly absorbed and exhibited excellent dose-proportionality with minimal between-subject variation in exposure. In poor sleepers, 40 µg of buccal DEX shortened the sleep latency by 11 min, increased the time spent in non-rapid-eye-movement sleep by 38 min, and elevated electroencephalographic slow wave energy (0.75-4.0 Hz) in the first half of the night by 23 % (P_all_ < 0.05). Rapid-eye-movement sleep latency was dose-dependently prolonged (20 µg: 48 min; 40 µg: 117 min; P_all_ < 0.01). Nocturnal cortisol, melatonin and heart rate, and morning cortisol were not significantly affected by DEX, nor did post-awakening orthostatic regulation, subjective sleepiness and mood, and psychomotor vigilance differ among the conditions.

**Conclusions:** The favorable PK-PD profile of oro-mucosal DEX delivery warrants further dose-finding and clinical studies, to establish the exact roles of α_2_ receptor agonism in pharmacological sleep enhancement and as possible novel mechanism to alleviate stress-related insomnia.

## INTRODUCTION

Insomnia is the second most prevalent neuropsychiatric disorder. Although providing symptomatic relief, currently approved sleep medications do not promote physiological sleep processes and are prone of unwanted effects like dependence, tolerance, and daytime impairments^1,2^ Thus, there exists an unmet clinical need for novel pharmacotherapeutic approaches to restore physiological sleep functions.

The locus coeruleus-noradrenergic (LC-NE) system may be a promising new target to ameliorate stress-induced sleep disturbances^3^ The LC-NE represents the main source of brain norepinephrine (NE) synthesis and constitutes an integral part of the ascending reticular activating system^4^. By acting on specific a and adrenoceptors, the LC-NE system also regulates essential waking functions including attention, arousal, memory, emotions, and cognition. The LC-NE activity is high in wakefulness, lower during non-rapid-eye-movement (NREM) sleep, and reaches near complete silence in rapid-eye-movement (REM) sleep^5^. Because uniquely low levels of NE are reached in REM sleep, consolidated REM sleep may be beneficial to normalize stress and negative affect across the night^3,6^ Thus, it has been hypothesized that stable REM sleep optimally favors adaptive synaptic plasticity during replay of emotional memory traces in the limbic circuit, which is highly active in this sleep stage. Entering sleep with lingering arousal prevents LC-NE shut-off required for sound sleep, and sustained LC-NE activity not only fragments REM sleep but also reduces deep NREM sleep. Such sleep disturbances are seen in many neuropsychiatric patients^7,8^ In addition, restless sleep may not merely be a symptom of an underlying neuropsychiatric disease, but may itself play a crucial role in the pathogenesis and progression of the disorder. Given these mutual relationships, reducing LC-NE activity could provide a promising strategy to break the vicious circle of disturbed nocturnal sleep and affective disorders.

The highly selective α_2_ adrenoceptor agonist, dexmedetomidine (DEX), recently elicited interest as potential repurposing candidate to enhance sleep in various medical conditions^9,10^. DEX activates inhibitory, presynaptic α_2_ receptors on the LC and inhibits the release of NE^11–13^. The compound is registered as intra-venous *(i.v.)* anesthetic and widely used to optimize sedation and analgesia during surgery, with reduced risk of post-operative delirium^14^. In patients undergoing emergency trauma surgery, low-dose DEX prevented the development of post-surgery PTSD symptoms and improved overall sleep quality^15^. Furthermore, *i.v.* administration of DEX improved sleep quality in patients with severe treatment-resistant insomnia^16^.

Because of pronounced first-pass metabolism^17,18^ DEX administration currently requires *i.v.* injection in controlled settings. These restraints hamper the possible use of DEX as sleep medication at home. Prior attempts at non-invasive (e.g., oral, nasal) DEX delivery were characterized by low and highly variable absorption^18,19^ To overcome these challenges, we developed innovative, fast­ disintegrating, oro-mucosal formulations for sublingual or buccal DEX administration. To establish the feasibility of oro-mucosal DEX delivery, we conducted two randomized, controlled trials and comprehensively tested the pharmacokinetics and sleep effects of two sub-anesthetic (20 & 40 µg) DEX doses in healthy male good sleepers and poor sleepers.

## MATERIALS AND METHODS

### Permission

The study was first submitted on 31.07.2020 for ethical approval by the Cantonal Ethics Committee of the Canton of Zurich (BASEC# 2020-016114; principal investigator: Oliver G. Bosch, MD). Upon receipt of the approval it was registered on ClinicalTrials.gov (Identifier# NCT04508166). All participants provided written informed consent according to the declaration of Helsinki.

### Study participants

The pharmacokinetic-pharmacodynamic (PK-PD) properties of oro-mucosal DEX were investigated in two separate studies. We conducted the first study in eight healthy young men with good sleep quality (referred to as “good sleepers”) and the second study in 17 healthy young men reporting subclinical insomnia (“poor sleepers”). Apart from the study populations, the two studies also slightly differed in the galenic formulations and the exact routes of oro-mucosal DEX application (see below). The following inclusion criteria were required for enrolment in both studies: male sex, to avoid unrecognized pregnancy and the impact of the menstrual cycle on sleep physiology or hypothalamus-pituitary axis activity; age range: 18-35 years; body-mass-index: 18.5-25 kg/m^2^; regular habitual sleep-wake rhythm (bedtime 22-24h); absence of somatic or psychiatric disorders; normal or corrected-to-normal vision; no acute or chronic medication intake; non-smoking; no history of drug abuse(> 5/lifetime; exception: occasional cannabis use); moderate alcohol(< 5 units/week) and caffeine consumption (< 4 units/day); no crossing of> 2 time zones within 30 days before study start. For inclusion in the second study, an Insomnia Severity Index (ISi) score between 8-14 was required.

Before final enrolment as study participant, all volunteers completed a screening night in the sleep laboratory, to exclude sleep-related disorders such as sleep apnea (Apnea-Hypopnea-lndex > 5/h of sleep), periodic limb movements(> 5/h) or sleep onset REM sleep episodes. Enrolled participants were required to abstain from illicit drugs, starting two weeks prior to the first experimental night until the end of the study (the day after the third experimental night). Intake of alcohol was not allowed within 24 hours of the start of the experimental nights. Participants were also instructed to keep an individual, regular sleep-wake rhythm with eight hours time-in-bed during the entire study, starting two weeks prior to the first experimental night. The rhythm was chosen depending on the volunteers’ habitual bedtime. All included participants chose to keep a bedtime between 22:00-24:00 and a corresponding rise time between 6:00-8:00. On average, bedtime occurred at 23:24 ± 0.31 min. Adherence to the sleep-wake schedule was monitored with a rest-activity monitor (GENEActiv, Activinsights Ltd., Kimbolton, UK) worn on the wrist of the non-dominant arm and a sleep-wake diary. Fifteen min before bedtime, an evening questionnaire was completed daily throughout the entire study which, on experimental nights, also served the evaluation of the consumption of drugs, alcoholic or caffeinated beverages and the adherence to the regular sleep­ wake rhythm.

The demographic characteristics of the study participants are reported in Supplementary Table Sl.

### Study drugs

#### Sublingual ODT

For the study in the good sleepers’ group, a first sublingual oro-dispersible tablet (ODT) formulation was developed by direct tablet compaction of DEX-HCI with tricalcium phosphate as bulking agent and croscarmellose sodium as super-disintegrant^20^. For this purpose, Dexdor™ injectable solution (100 µg/ml; Orion Pharm AG, Zug, Switzerland) was added in a Rotavap flask to tricalcium phosphate powder (Galvita AG, 4058, Switzerland), and the mixture was dried under vacuum in a rotary evaporator (40° C; 2 hours). Ac-Di-Sol (3%; FMC) was added as super­ disintegrant, and the blend was homogenized for ten min in a turbula mixer. Then, 104.7 mg of the blend was filled into the 7 mm die and compacted with 1 mT. The placebo tablets were obtained accordingly, but Dexdor™ was replaced by saline 0.9% (B. Braun Medical AG, Sempach, Switzerland) to mimic the slight salty taste of the Dexdor™ solution. All tablets were stored at room temperature under dry conditions (desiccator bag).

#### Buccal ODT

For the study in the poor sleepers’ group, an improved buccal ODT formulation was developed by freeze-drying for buccal delivery. Dextran FP40 (SERVA Electrophoresis GmbH; 69115 Heidelberg; Germany) was used as bulking agent and dissolved in Dexdor™ injectable solution (100 µg/ml; Orion Pharm AG, Zug, Switzerland). The solution was then volumetrically filled into aluminum blister molds (0.2 ml/cavity) using an Eppendorf Micropipette and finally freeze-dried for 30 hours, to yield lyophilized tablets with a strength of 20 µg each. The placebo melting tablets were obtained accordingly, but Dexdor™ was replaced by saline 0.9% (B. Braun Medical AG, Sempach, Switzerland), to mimic the slight salty taste of the Dexdor™ solution. All tablets were stored at room temperature under dry conditions (desiccator bag).

We examined the PK-PD profiles of sublingual and buccal DEX administration. The first study in the good sleepers (sublingual ODT) mainly served as pilot study, to evaluate the performance of oro­ mucosal DEX delivery, identify an adequate dose range, and determine preliminary effects on sleep physiology.

### Study design

Both studies adhered to a randomized, placebo-controlled, balanced, double-blind, cross-over design. The study protocols consisted of one screening and three experimental nights (placebo, 20 and 40 µg of DEX) separated by a washout period of 7 days. The placebo and verum tablets were combined to yield total doses of either O µg (2 x placebo), 20 µg (1 x 20 µg + 1 x placebo), or 40 µg (2 x 20 µg) of DEX. The assessment of the compound’s effect on neurobehavioral, emotional, cognitive, and endocrinological markers of sleep inertia followed immediately after awakening. To simplify descriptions and data presentations, we will refer to the scheduled, 23:00-07:00 time-in-bed with respect to the time-points of the tasks, because most of the participants roughly followed this sleep­ wake schedule. The details of the study design are illustrated in Fig. lA.

### Bioanalyses of blood plasma samples

#### Blood sampling

After drug intake at bedtime, study participants were allowed to sleep for 8 hours in the soundproof and climatized bedrooms of the sleep laboratory. Blood samples were continuously collected from the left (in case of unsuccessful venipuncture from the right) antecubital vein via a venous catheter connected with Heidelberger plastic tube extensions to the adjacent room. Blood was drawn at baseline (30 min before drug intake) and 0.25, 0.5, 0.75, 1, 1.25, 1.5, 1.75, 2, 2.5, 3, 3.5, 4, 5, 6, 7, 8, and 9 h after drug administration (4 ml, BD Vacutainer EDTA), while keeping the disturbance of the participants’ sleep to a minimum. The peripheral venous access was kept patent via a constant drip (10 ml/h) of heparinized saline (1000 IU heparin in 0.9 g NaCl/di; HEPARIN Bichsel; Bichsel AG, 3800 Unterseen, Switzerland). Blood samples were centrifuged for five min at 2000 RCF immediately after blood drawing. The resulting plasma samples were directly transferred to –20° C until final storage at –80° C.

#### Quantification of dexmedetomidine

The DEX was purchased from Sigma-Aldrich (St. Louis, USA) and medetomidine-^13^C-d3was purchased from Cayman chemical (Ann Arbor, USA) and used as internal standard. All chemicals used were of the highest purification grade available. Six hundred µI of plasma and 30 µI of IS (8 ng/ml) were added to a tube. For calibrator and quality control samples, an additional 100 µI of the calibrator or quality control solutions were added. A liquid-liquid extraction using 1000 µI of ethyl acetate/butyl acetate (1:1 v/v) was performed by shaking the tubes for 10 min and centrifuging them at 12’000 rpm for 5 min. Nine hundred µI of the organic phase were transferred into a vial and evaporated to dryness under a gentle stream of nitrogen, before adding 50 µI of an eluent-mixture (85:15 v/v) for reconstitution. The samples were analyzed on an ultra-high performance liquid chromatography (UHPLC) system (Thermo Fisher, San Jose, CA), coupled to a linear ion trap quadrupole mass spectrometer 5500 (Sciex, Darmstadt, Germany). The mass spectrometer was operated in positive electrospray ionization mode with multiple reaction monitoring. Three transitions were used for DEX: 200.9 95.3, 200.9 68, 200.9 41. The mobile phases of the UHPLC system consisted of water (eluent A) and acetonitrile (eluent B), both containing 0.1 % of formic acid (v/v). Two µI of the samples were injected using a Kinetex C18 column (100 x 2.1 mm, 1.7 µm) (Phenomenex, Aschaffenburg) with the flow rate set to 0.5 ml/min. The flow gradient started with 85 % of eluent A for 0.5 min, decreasing to 50 % within 1 min, and increasing back to 85 % of eluent A in 0.5 min. These conditions were held for 1 min. For quantification, the peak area of DEX was further integrated and divided by the peak area of the internal standard. Calibrator samples were fitted with a least-squares fit and weighted by 1/x. The limit of quantification of DEX was 0.01 ng/ml and the limit of detection was 0.005 ng/ml.

#### Quantification of cortisol

Plasma cortisol levels were measured in blood samples collected at baseline, 90, 180, 360, and 480 min after drug administration. The plasma samples were spiked with 38 µI IS, 250 µI ZnSO4 was added and 500 µI of purified water. They were vortexed and centrifuged at 8000 g for 5 min. Afterwards, the samples were purified using a solid-phase extraction on an OasisPrime HLB 96-well plate using a positive pressure 96-well processor (both Waters, UK). For LC­ MS analysis, a Vanquish UHPLC (equipped with an ACQUITY UPLC HSS T3 Column, 100A, 1.8 µm, 1 mm X 100 mm column; Waters, Switzerland) was coupled to a Q Exactive Plus Orbitrap (both Thermo Fisher Scientific, Reinach, Switzerland). Separation was achieved using gradient elution over 11 min using water and methanol both supplemented with 0.1 % formic acid (all Sigma-Aldrich, Buchs, Switzerland) as mobile phases. Data analysis was performed using TraceFinder 4.1 (Thermo Fisher Scientific, Reinach, Switzerland). The method was validated according to international standards (Andrieu et al, 2022). Steroid hormone concentrations were calculated in nmol/1.

#### Quantification of melatonin

Plasma melatonin levels were measured in blood samples collected at baseline, 90, 180, 360, and 480 min after drug administration. Melatonin was quantified with LC­ MS/MS, as previously reported elsewhere (Dornbierer *et al.,* 2019).

### Polysomnography

Sleep was recorded by all-night polysomnography from 23:00 to 07:00 with dedicated polysomnographic amplifiers (SIENNA ULTIMATE®, EMS Biomedical, Korneuburg, Austria). The recording montage consisted of 14 EEG electrodes according to the 10-20 system (Fpl, Fp2, F3, F4, Fz, C3, C4, Cz, P3, P4, Pz, 01, 02, Oz), a bipolar electrooculogram (EOG), a submental electromyogram (EMG), and a two-lead electrocardiogram (ECG). The analogue signals were conditioned by a high-pass filter (EEG: 0.5 Hz; EMG: 5 Hz; ECG: 1 Hz) and a low-pass filter (EEG: 70 Hz; EMG: 100 Hz; ECG: 70 Hz), digitized and stored at the sampling frequency of 512 Hz.

#### Visual scoring

Sleep variables were visually scored based on 30-s epochs according to the criteria of the American Academy of Sleep Medicine^21^. For sleep scoring, the F3-A2, C3-A2 and O1-A2 (in case of severe artifacts: F4-Al, C4-Al, O2-Al) derivations were used. Movement– and arousal-related artifacts were visually identified and excluded from the analyses. The following sleep variables were analyzed: time-in-bed: time between lights-off and lights-on; total sleep time: time spent in stages Nl, N2, N3, and REM sleep; sleep efficiency: percentage of total sleep time per time-in-bed; sleep latency: time between lights-off and first occurrence of three consecutive epochs of Nl sleep or any other sleep stage; REM sleep latency: time between sleep onset and the first occurrence of REM sleep; time spent in wakefulness after sleep onset, NREM sleep stages Nl, N2, N3, and REM sleep.

#### Quantitative EEG analysis

Following notch filtering, a low-pass filter with a cutoff frequency of 40 Hz and high-pass filter with a cutoff frequency of 0.5 Hz were applied. The EEG signal was then down-sampled to 128 Hz and C3 and C4 electrodes were re-referenced to the mastoids (C3-M2 and C4-Ml). The filtered signal was decomposed in the frequency domain with the Welch’s averaged modified periodogram, using a 4-s Hanning window with 50 % overlap, resulting in a 0.25-Hz frequency resolution. The EEG spectral power between 0.75-4.0 Hz was calculated for the derivations C3-M2 and C4-Ml. The derivation with the better signal quality was used for averaging. Furthermore, EEG slow wave energy in the first and second half of the night was computed as the cumulative sum of delta power (0.75-4.0 Hz) across artifact-free N2 and N3 sleep episodes. The EEG pre-processing and computational analyses were carried out in MATLAB (R2024a, The Math-Works, Inc., Natick, Massachusetts, USA).

### State upon awakening

#### Cortisol Awakening Response

To quantify the cortisol awakening response, we sampled saliva in each participant at 07:00 (immediately after awakening), and at 07:15, 07:30, 07:45, 08:00 and 08:15. Participants were instructed to chew a cotton swab for 60 sand return it into the dedicated Salivette® tube (Sarstedt, Germany). The tubes were immediately stored on ice until final storage at –80° C. Cortisol was quantified with LC-MS/MS, as reported elsewhere^22^.

#### Orthostatic regulation

We conducted the Schellong test, *aka* Scheilong maneuver, immediately after awakening (7:00), to assess potential carry-over effects of DEX on post-awakening orthostatic regulation^23^. This validated clinical test involves changing from a supine to a standing body position and monitoring various physiological variables, including blood pressure and heart rate. The blood pressure and heart rate were measured at one-minute intervals, first three times in a supine and then five times in a standing position. Because of measurement errors, two assessments in the placebo condition and one assessment in the 20 µg DEX condition needed to be excluded from the analyses. In addition, one assessment in the 20 µg DEX condition was discontinued because the participant felt dizzy. To identify orthostatic dysregulations, the difference between the last blood pressure measurement in the supine position and the first three measurements in the upright position were calculated. A drop of 20 mm Hg in systolic and/or 10mm Hg in diastolic blood pressure are considered orthostatic hypotension events^24^. To document the mean blood pressure and heart rate values during the Scheilong test, means across all time points per position were calculated.

#### Morning Sleep Questionnaire

We used a morning sleep questionnaire^25^, to assess subjective sleep quality, including estimated time to fall asleep, number of nocturnal awakenings, wake time after sleep onset, and visual analogue scales to compare subjective sleep quality against a habitual night at home.

#### Sleep Inertia Questionnaire

The sleep inertia questionnaire^26^ was designed as a trait inventory, where participants are instructed to rate the quality of their awakening over the past week, focusing on physiological, emotional, cognitive, and behavioral aspects. The instructions for the inventory were as follows: ‘On a typical morning in the past week, after you woke up, to what extent did you, for example, have trouble getting out of bed?’ (with possible ratings ranging from l=’not at all’ to S=’all the time’).

For the current study, we adapted the original sleep inertia questionnaire to assess the acute subjective experiences of volunteers at waking up^22^. We rephrased the inventory instructions to gather information about the state of the waking process on the experimental morning, rather than trait information about the previous week. This allowed us to analyze possible immediate effects of the study drug. The revised instructions read as follows: ‘How strongly did you experience the following aspects after waking up this morning, in comparison to a typical morning last week, such as having trouble getting out of bed?’ (with possible ratings from –3=’extremely less’ to 3=’extremely more’). For our purposes, the questionnaire was renamed as Acute Sleep Inertia Questionnaire and administered at 07:45.

#### Subjective sleepiness

Before (at 22:40) and after (at 07:15) each experimental night, we administered the validated Karolinska Sleepiness Scale^27^.

#### Positive and Negative Affect Schedule (PANAS)

We administered the PANAS (Watson et al, 1988) at 07:30, a questionnaire in which the participant is asked to rate the occurrence and intensity of 20 mood states (ten positive and ten negative adjectives) on a 5-point Likert scale at the moment of the rating.

#### State-Trait Anxiety Inventory

We used the State-Trait Anxiety Inventory^28^, to assess the participants’ trait anxiety level at the screening night and the state anxiety symptoms at 07:35 of every experimental morning. Both questionnaires consist of 20 items which are rated on 4-point Likert-scales.

#### Psychomotor Vigilance Test

Thirty min before bedtime/drug intake and 15 min after awakening, we administered a 10-min version ofthe psychomotor vigilance test^29^ and analyzed the median reaction time (RT) and numbers of lapses (trials with RT> 500 ms).

### Statistical analyses

We computed independent linear mixed-effects models, with *’condition’* (placebo, 20 µg DEX, 40 µg DEX), *’time’* (cortisol awakening response: 07:00, 07:15, 07:30, 07:45, 08:00, and 08:15; psychomotor vigilance test: 22:30 and 07:15) and *’body position’* (supine, upright) as within­ participant factors, and *’participant ID’* as random effect. We employed ‘R’ for all analyses (R Versions 4.3.1 and 4.3.2; RStudio, Inc.; R-package “lme4,” Version 1.1-34). In all applied models, we applied normal Q-Q plots to demonstrate normality of the residuals. Moreover, we verified the assumption of homoscedasticity and linearity using a Tukey-Anscombe plot (residuals vs. fitted).

When appropriate, we carried out post-hoc testing using the ‘R’ package emmeans (Version 1.8.9). We corrected the p values of the post-hoc tests for multiple comparisons using Benjamini-Hochberg correction of the false discovery rate.

## RESULTS

### Pharmacokinetics of oro-mucosal dexmedetomidine intake

As illustrated in Figs. 1B & lC, the pharmacokinetic profiles of both oro-dispersible formulations were exceptionally stable. The DEX plasma concentration data were modeled by a non­ compartmental, first-order absorption, and first-order elimination pharmacokinetic model. The pharmacokinetic features of the two formulations are summarized in Table 1.

**Figure 1:**
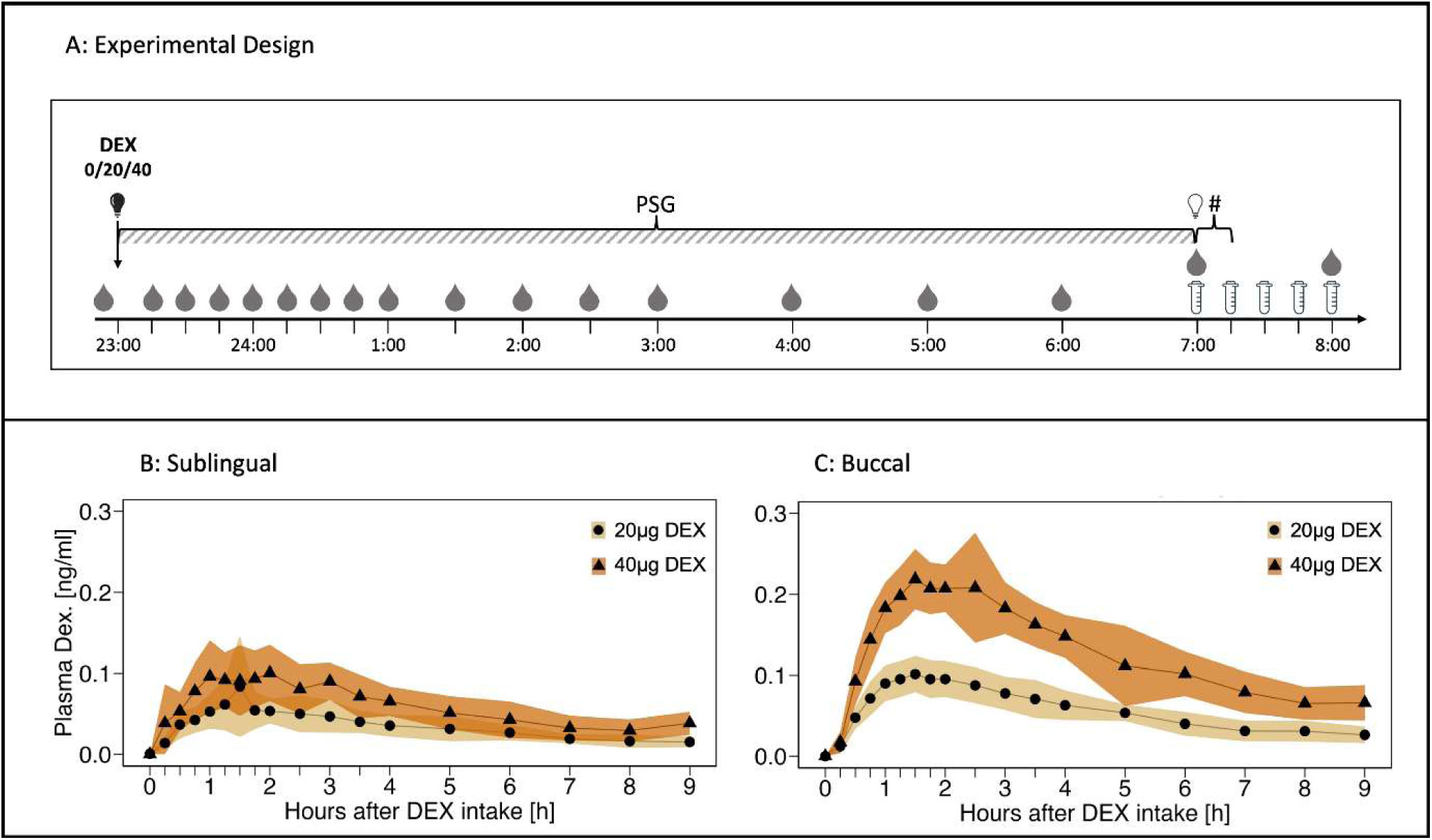
Experimental design and pharmacokinetic profiles of dexmedetomidine (DEX) after sublingual and buccal oro-dispersible tablet (ODT) intake. **(A)** Time points of blood draws (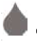 and tick marks on x-axes), saliva sampling (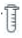), and post-awakening testing(#) are indicated. The study drugs (0/20/40 µg DEX) were administered at 23:00 when lights were switched off (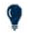) and the polysomnographic recording was initiated (time point ‘0’). The participants were awoken and lights were turned on at 07:00 (time point ‘8’). **(B)** Evolution of DEX plasma concentration after intake of sublingual ODT (n = 8). **(C)** Evolution of DEX plasma concentration after intake of buccal ODT (n = 17). Yellow lines connecting black dots: 20 µg DEX intake at bedtime. Orange lines connecting black triangles: 40 µg DEX intake at bedtime. Yellow/orange shadings indicate standard deviations.

**Table 1.**
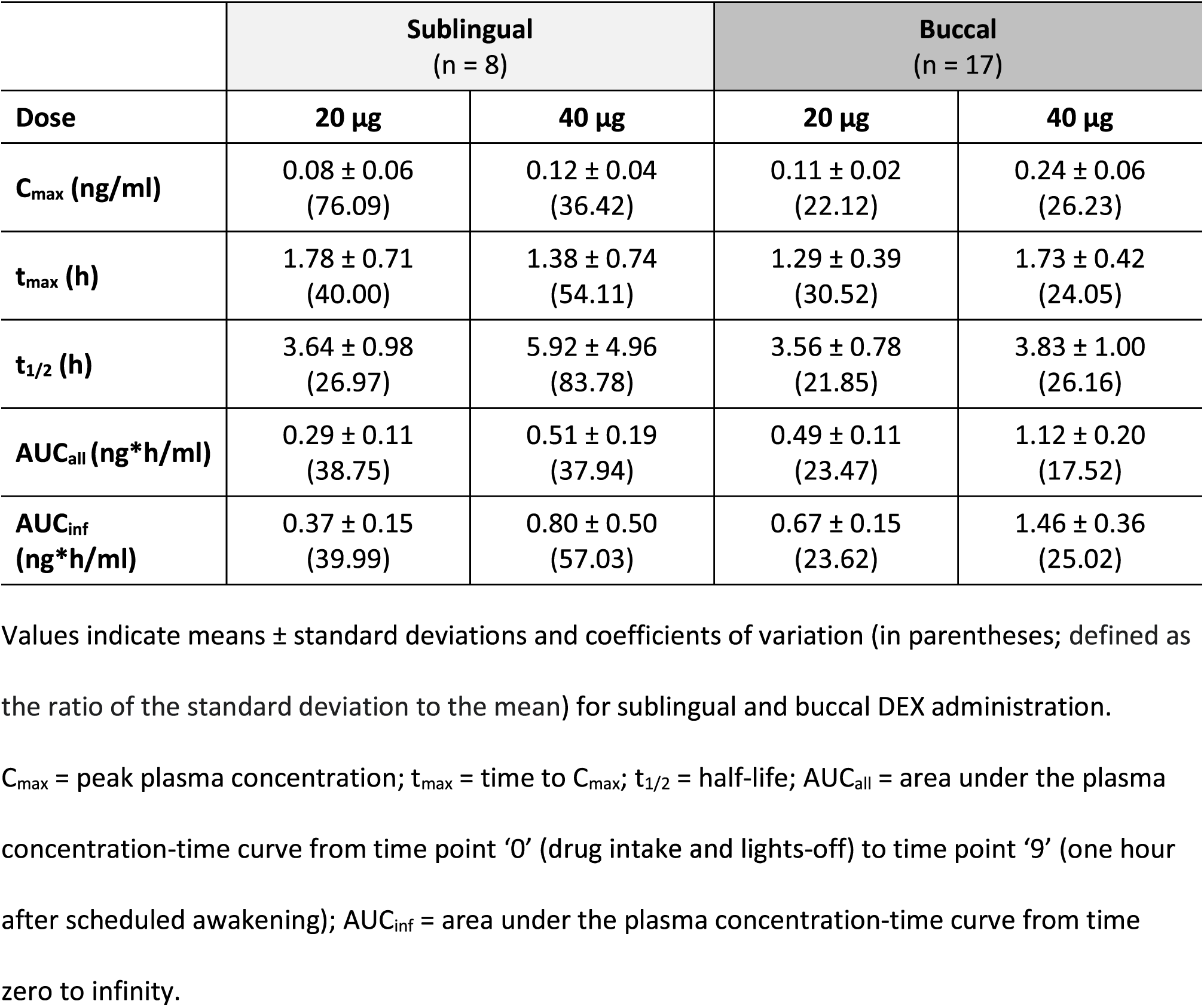
Pharmacokinetic parameter estimates during sleep after sublingual and buccal DEX intake.

|  | Sublingual<br>(n = 8) |  | Buccal<br>(n = 17) |  |
| --- | --- | --- | --- | --- |
| Dose | 20 µg | 40 µg | 20 µg | 40 µg |
| **C<sub>max</sub> (ng/ml)** | 0.08 ± 0.06<br>(76.09) | 0.12 ± 0.04<br>(36.42) | 0.11 ± 0.02<br>(22.12) | 0.24 ± 0.06<br>(26.23) |
| **t<sub>max</sub> (h)** | 1.78 ± 0.71<br>(40.00) | 1.38 ± 0.74<br>(54.11) | 1.29 ± 0.39<br>(30.52) | 1.73 ± 0.42<br>(24.05) |
| **t<sub>1/2</sub> (h)** | 3.64 ± 0.98<br>(26.97) | 5.92 ± 4.96<br>(83.78) | 3.56 ± 0.78<br>(21.85) | 3.83 ± 1.00<br>(26.16) |
| **AUC<sub>all</sub> (ng\*h/ml)** | 0.29 ± 0.11<br>(38.75) | 0.51 ± 0.19<br>(37.94) | 0.49 ± 0.11<br>(23.47) | 1.12 ± 0.20<br>(17.52) |
| **AUC<sub>inf</sub>**<br>**(ng\*h/ml)** | 0.37 ± 0.15<br>(39.99) | 0.80 ± 0.50<br>(57.03) | 0.67 ± 0.15<br>(23.62) | 1.46 ± 0.36<br>(25.02) |

Values indicate means ± standard deviations and coefficients of variation (in parentheses; defined as the ratio of the standard deviation to the mean) for sublingual and buccal DEX administration.
C<sub>max</sub> = peak plasma concentration; t<sub>max</sub> = time to C<sub>max</sub>; t<sub>1/2</sub> = half-life; AUC<sub>all</sub> = area under the plasma concentration-time curve from time point '0' (drug intake and lights-off) to time point '9' (one hour after scheduled awakening); AUC<sub>inf</sub> = area under the plasma concentration-time curve from time zero to infinity.

The sublingual formulation (Fig. 1B) reached maximum plasma concentrations of 0.08 ± 0.06 ng/ml within 1.78 ± 0.71 h for the 20 µg dose and 0.12 ± 0.04 ng/ml within 1.38 ± 0.74 h for the 40 µg dose. The estimated terminal plasma half-lives were 3.64 ± 0.98 hand 5.92 ± 4.96 h, and the AUC were 0.29 ± 0.11 ng*h*mI-^1^ and 0.51 ± 0.19 ng*h*mI-^1^for the 20 and 40 µg doses.

The corresponding characteristics for the 20 and 40 µg doses of the improved buccal formulation (Fig. lC) were: maximal plasma concentrations of 0.11 ± 0.02 ng/ml and 0.24 ± 0.06 ng/ml within 1.29 ± 0.39 hand 1.73 ± 0.42 h. The estimated terminal plasma half-lives were 3.56 ± 0.78 hand 3.83 ± 1.00 hand the AUC were 0.49 ± 0.11 ng*h*mI-^1^ and 1.12 ± 0.20 ng*h*mI-^1^. All pharmacokinetic variables exhibited an exceptionally low interindividual variation.

### Sleep variables

#### Sublingual ODT

Compared to placebo, both the 20 and 40 µg doses of DEX prolonged the time spent in NREM sleep (combined stages N2 & N3) and delayed the latency to REM sleep (Fig. 2). Both doses tended to increase the time spent in N2 sleep (p:’> 0.1), whereas the 40 µg dose tended to reduce the duration of REM sleep (p < 0.07). All other polysomnographic sleep variables remained unaffected by the drug (Supplementary Table S2).

**Figure 2.**
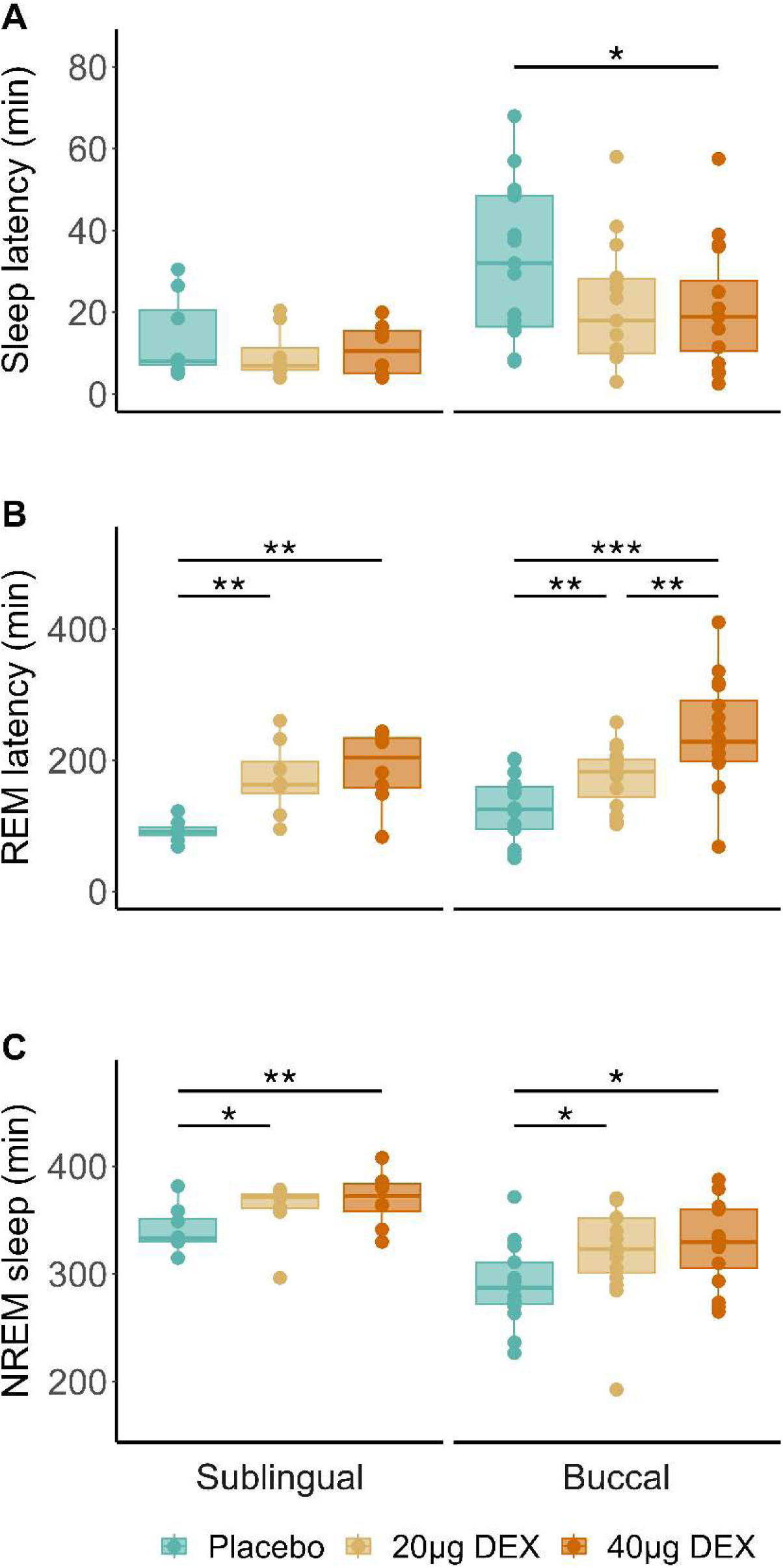
Visually scored sleep variables following intake of sublingual (left panels; n = 8) and buccal (right panels; n = 17) oro-mucosal DEX formulations at bedtime. **(A)** Sleep latency until three consecutive epochs of Nl or deeper sleep stages. **(B)** Latency to first REM sleep episode. (C) Total duration of NREM sleep stages N2 and N3. Box plots: horizontal lines mark the median, lower and upper hinges correspond to the 25^th^ and 75^th^ percentiles, and whiskers extend to the last value within 1.Sx of the interquartile range. Dots: individual data points. Blue: placebo; yellow: 20 µg DEX; orange: 40 µg DEX. Statistically significant differences between conditions are denoted by asterisks. *\*p < 0.05; **p < 0.01; ***p < 0.001 (Benjamini-Hochberg corrected)*

#### Buccal ODT

Compared to placebo, both the 20 (p = 0.053) and 40 µg doses of DEX shortened sleep latency by roughly 11 min (Fig. 2). Moreover, both doses prolonged the time spent in N2 sleep, as well as the combined stages N2 & N3. The DEX dose-dependently delayed the occurrence of REM sleep, while the higher dose tended to reduce REM sleep duration (p = 0.071). All other visually scored sleep variables remained unaffected by the DEX when compared to placebo (Supplementary Table S2).

Upon confirmation that both oro-mucosal DEX formulations similarly affected sleep architecture in good and poor sleepers, the subsequent analyses were restricted to the pharmacokinetically superior buccal formulation, which was studied in the poor sleepers.

### Quantitative analysis of the EEG in NREM sleep

In all experimental nights, EEG slow wave energy in NREM sleep decreased from the first to the second half of the sleep episodes (Pa11<0.0001), reflecting the dissipation of sleep pressure (Fig. 3). Compared to placebo, 40 µg DEX increased slow wave energy in NREM sleep in the first four hours of the sleep episode (7.07 ± 3.94 *vs.* 5.45 ± 3.65 µV^2^/Hz x 10^4^; p < 0.03). After intake of 20 µg DEX, slow wave energy tended to be higher than after placebo (6.26 ± 3.15 *vs.* 5.45 ± 3.65 µV^2^/Hz x 10^4^; p = 0.055). No differences among the three experimental conditions were observed in the second half of the night.

**Figure 3.**
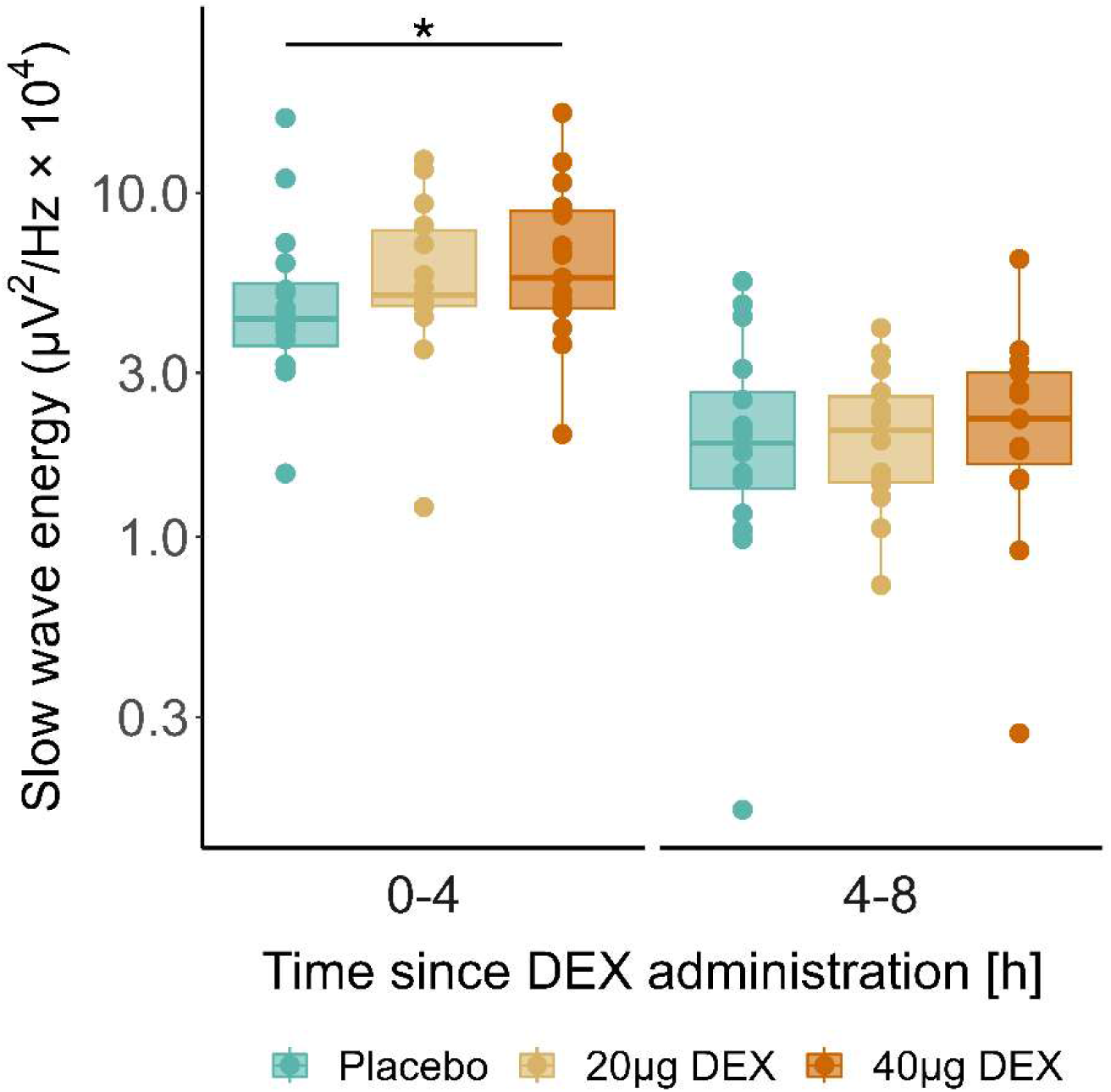
The EEG slow wave energy in the 0.75-4.0 Hz range in NREM sleep stages N2 & N3 was quantified in the first (0-4 h after intake) and second (4-8 h after intake) halves of the nights following buccal DEX administration at bedtime. Box plots: horizontal lines mark the median, lower and upper hinges that correspond to the 25^th^ and 75^th^ percentiles, and whiskers extend to the last value within 1.Sx of the interquartile range. Dots: individual data points (n = 17). Blue: placebo; yellow: 20 µg DEX; orange: 40 µg DEX. Note that the values are presented on a logarithmic scale. The asterisk indicates the significant difference between the 40 µg DEX and placebo conditions in the first half of the experimental nights. Note that data are plotted on a logarithmic scale. * p < 0.03 (Benjamini-Hochberg corrected)

### Endocrinological and cardiovascular assessments

Neither the mean nocturnal blood plasma cortisol concentrations, the nocturnal plasma melatonin profiles, nor the mean cortisol level in saliva during assessment of the cortisol awakening response did differ between the placebo and the drug conditions (Table 2). Furthermore, no differences in mean nocturnal heart rate in NREM and REM sleep were detected. The Scheilong Test conducted upon awakening identified very few orthostatic hypotension events that did not differ among the experimental conditions (placebo: n=0/15; 20 µg DEX: n = 2/15; 40 µg DEX: n = 1/17). Heart rate, as well as mean systolic and diastolic blood pressure in supine and upright positions revealed a normal orthostatic response and did not differ among the conditions (Table 2).

**Table 2.**
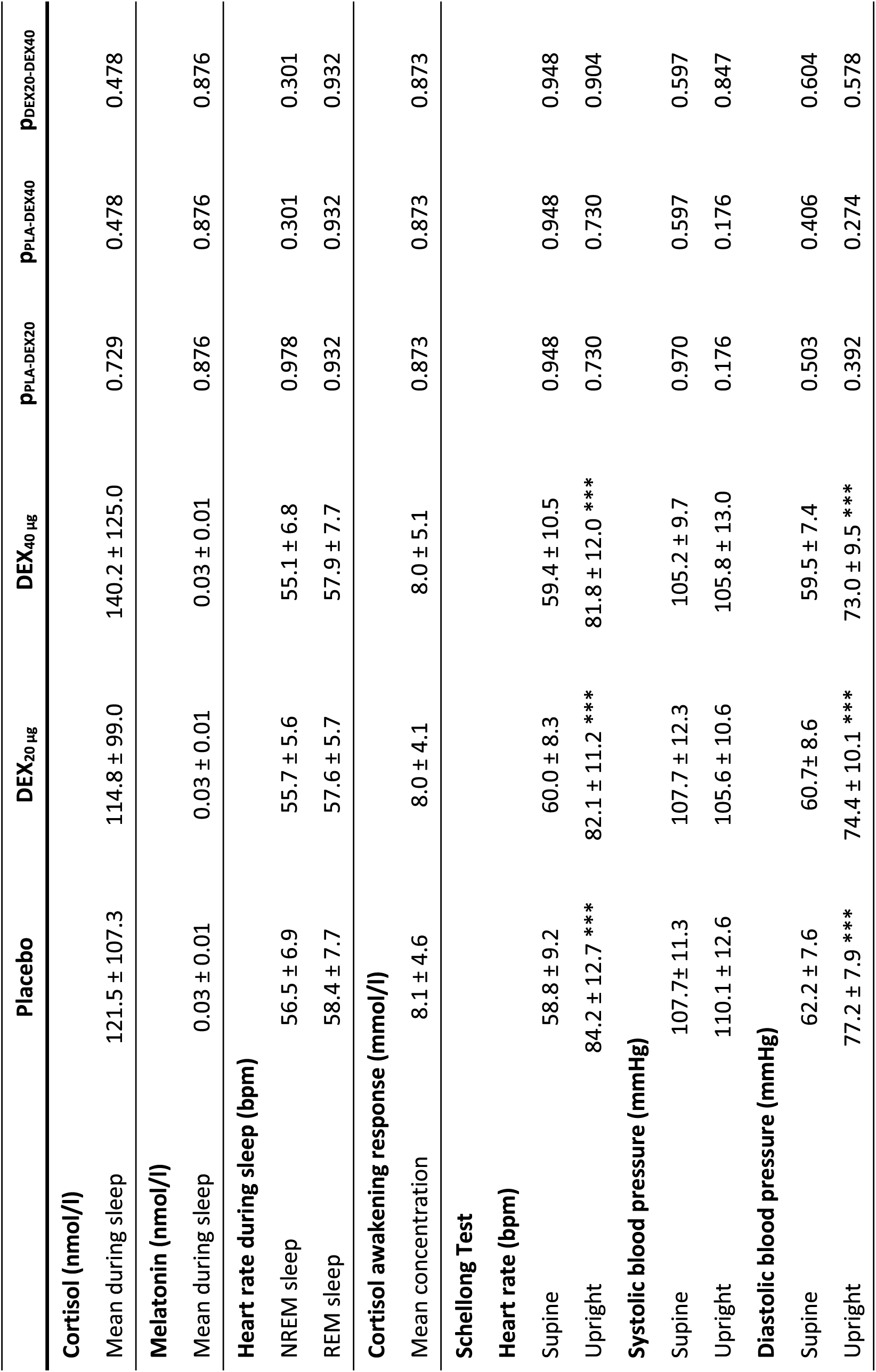
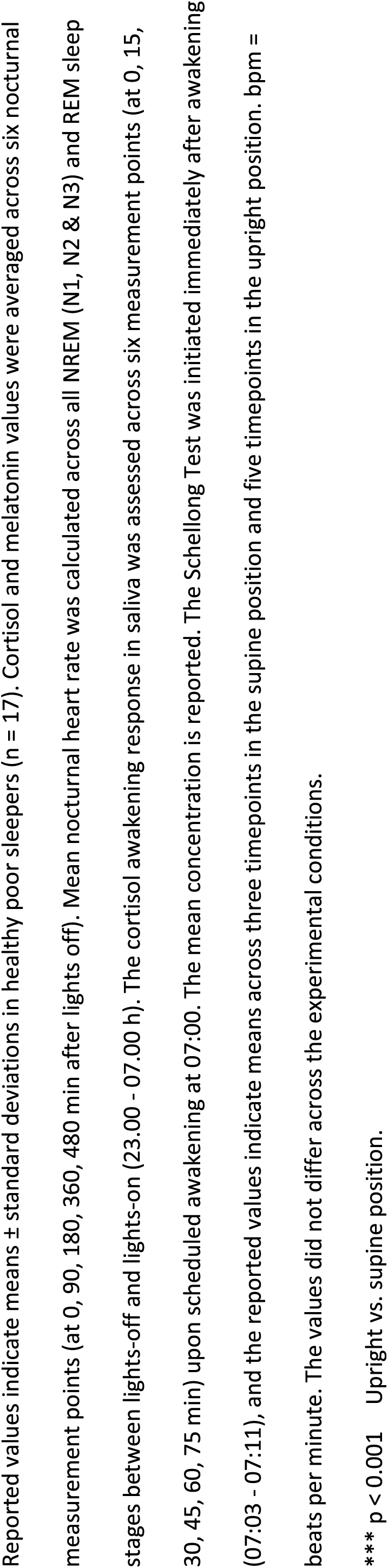
Endocrinological and cardiovascular assessments during sleep and upon awakening.

### Subjective sleep quality, self-rated state and vigilance upon awakening

Validated questionnaires and neurobehavioral tests were employed to assess subjective sleep quality, self-rated state, and neurobehavioral vigilance upon scheduled awakening. The statistical analysis revealed no differences among the experimental conditions. The data are summarized in Supplementary Table S3.

## DISCUSSION

Despite accumulating preclinical and clinical evidence supporting the repurposing potential of DEX to ameliorate stress-related sleep disturbances^10^, no DEX delivery system is currently available for insomnia management in outpatient settings. With this purpose in mind, we developed two oro­ mucosal, fast-disintegrating DEX formulations (one sublingual and one buccal) and assessed their clinical potential to enhance sleep quality at sub-anesthetic (20 & 40 µg) doses. The pharmacokinetic analyses during sleep revealed exceptional pharmaceutical properties, particularly of buccal delivery, characterized by rapid absorption, excellent dose-proportionality, and minimal variability in exposure among the study participants. The polysomnographic analyses further demonstrated the almost immediate onset of pharmacological effects, including markedly shortened sleep latency, prolonged time spent in NREM sleep, and delayed occurrence of REM sleep. The higher dose (40 µg) of buccal DEX administration also increased EEG slow-wave energy in the first half of the night. Importantly, neither of the very low doses administered affected cortisol, melatonin and heart rate during sleep, nor altered cortisol secretion, cardiovascular functions, as well as self-rated and neurobehavioral vigilance upon awakening.

Besides i.v.-injectable solutions used in anesthesia (USA: Precedex®; Europe: Dexdor®), the FDA recently approved a mucoadhesive film containing 120-180 µg DEX (lgalmi®) to ameliorate agitation in in-patients suffering from bipolar disorder and schizophrenia. Indeed, evidence accumulates that oro-mucosal routes of DEX administration exhibit reduced first-pass metabolism and lower inter­ subject variability in plasma concentrations than other non-invasive approaches such as oral and intra-nasal delivery^17–19^ Here we demonstrate in young men that between two slightly different oro­ mucosal DEX formulations, buccal administration showed approximately 50% higher bioavailability, considerably lower inter-subject variability in plasma exposure (coefficient of variation:∼ 20 vs.∼ 38), better dose-proportionality, and faster onset of action than sublingual administration. We suggest that the two systems not only differ in the mucoadhesive properties of the excipients, but also in saliva production and ensuing swallowing and first-pass degradation. Thus, we conclude that buccal delivery of sub-anesthetic DEX doses ensures an exceptional pharmacokinetic profile that offers a precise and convenient option for out-patient administration.

To assess the effects of 20 and 40 µg DEX on sleep, we studied two groups of healthy individuals reporting good and poor subjective sleep quality. The poor sleepers presented with subclinical insomnia (8 < ISi < 14), but did not meet the recommended ISi threshold for clinical insomnia disorder (ISi > 14). Nevertheless, they exhibited prolonged sleep latency, longer wake time after sleep onset, reduced sleep efficiency, and shorter NREM (stages N2 & N3) sleep duration than the good sleepers (Supplementary Table S2). Both doses of DEX hastened the onset of sleep in the poor sleepers, demonstrating a very rapid onset of action of buccal DEX intake at bedtime. Looking at the pharmacokinetic profiles of the two studies (Fig. 1), it appears that DEX plasma levels as low as 0.05 ng/ml can promote sleep. By contrast, sleep onset remained unchanged in the good sleepers. Given their short sleep latency in the placebo condition, they probably initiated sleep in the verum nights before a relevant DEX exposure level was present. In addition, the plasma concentration increased slower and reached a lower maximum with the sublingual formulation than with the buccal formulation. This difference may further explain the lack of sleep onset effects in the good sleepers.

Of note, we observed a pronounced and dose-dependent prolongation of REM sleep latency, which is consistent with previous studies^30,31^ The prolonged and more consolidated NREM sleep characterized by increased EEG slow wave energy in the first half of the night may have contributed to this effect. Given that a fixed sleep schedule with constant time-in-bed (eight hours) was employed, the delayed onset of REM sleep may have contributed to the marginally reduced REM sleep duration after both sublingual and buccal 40 µg DEX delivery (Supplementary Table S2). We suggest to provide the participants in future studies with a prolonged or *ad libitum* sleep opportunity, to address the question whether sub-anesthetic DEX administration only delays or also reduces the expression of REM sleep.

Interestingly, EEG slow wave energy was unchanged in the second half of the night, even though the DEX plasma levels were still in a concentration range (0.075-0.15 ng/ml) that enhanced slow NREM sleep oscillations in the initial hours of sleep, particularly after intake of 40 µg DEX. At the doses tested, DEX may consolidate deep NREM sleep at elevated homeostatic NREM sleep pressure rather than after the dissipation of sleep propensity. In addition, the effects of DEX on the sleep EEG may depend on the time of day of drug administration.

Further supporting this notion, subjective sleepiness was unaffected upon awakening in the morning, although a residual DEX concentration between 0.05-0.1 ng/ml was still present in plasma. Conversely, DEX plasma levels between 0.025-0.05 ng/ml shortened sleep latency at bedtime. Again, this finding may indicate that DEX facilitates sleep when sleep pressure is high, rather than inducing a mere pharmacological sedation. According to its mode of action, DEX inhibits the activity of wake­ promoting neurotransmitter systems, particularly LC-NE and histaminergic activity originating in the tuberomammillary nucleus^13,32,33^. These systems are disinhibited with decreasing homeostatic sleep drive^4,34^. Thus, when endogenous sleep propensity is low and endogenous circadian phase favors wakefulness, or in patients with elevated sympathetic activity such as during acute psychological agitation, a higher dose of DEX than the very low doses tested in this study may be necessary to promote sleep. In addition, we propose that buccal DEX could be very valuable in the perioperative setting because it provide effective sedation and anxiolysis with a lower risk of respiratory depression than traditional sedatives. Its convenient administration shown here could improve patient comfort and compliance, particularly in premedication and post-operative recovery.

### Limitations

Some limitations need to be considered when evaluating the results of the present studies. First, the study samples were rather small and homogenous, consisting exclusively of healthy, young, average­ weight Caucasian men. This precludes the generalization of the findings. Larger and more diverse samples are needed, to better ascertain the safety, tolerability, and efficacy of this intervention in clinical populations. Second, the effects of the two DEX formulations may not be perfectly comparable because the sublingual formulation was tested in good sleepers, while the buccal administration was tested in poor sleepers. We cannot be completely sure that the differences in the pharmacokinetic profiles are due to the different study samples, the slightly different oro-mucosal formulations, or a combination of these differences. Finally, to estimate the absolute bioavailability of the two formulations, *i.*v.-DEX delivery would be necessary.

### Conclusions

We developed two oro-dispersible galenic formulations containing 20 and 40 µg of the sympatholytic, α_2_ receptor agonist DEX and show that particularly buccal delivery has exceptional pharmacokinetic properties and exerts a very fast onset of action to promote and enhance NREM sleep in healthy poor sleepers. The intervention was safe, and we observed no orthostatic dysregulation, blunted cortisol awakening response nor impaired vigilance upon awakening. We conclude that buccal DEX is a unique tool to elucidate the roles of the LC-NE system in human sleep­ wake regulation. Furthermore, we suggest that clinical studies are warranted to establish α_2_ receptor agonism as a novel mode of action to treat stress-related insomnia.

## TRIAL REGISTRATION

ClinicalTrials.gov identification number: NCT04508166

## PUBLICATION HISTORY

This manuscript was previously posted to medRxiv (MS ID#: MEDRXIV/2024/309892).

## PRIOR PRESENTATIONS

Data of the paper were in part presented at the following conferences:

1) European Sleep Research Society Congress, 26.9.-30.9.2022, Athens, Greece
2) SLEEP 2023, American Academy of Sleep Medicine Congress, 3.6.-7.6.2023, Indianapolis, USA
3) World Sleep Conference, World Sleep Society, 20.10.-25.10.2023, Rio de Janeiro, Brazil

## Data Availability

All data produced in the present study are available after peer-reviewed publication upon reasonable request to the authors.

## ACKNOWLEDGEMENTS

We thank Dr. Ian Clark, Mr. Fabio Carbone, Ms. Tamara Hurlimann and Mr. Andreas Maag for their dedicated support in the preparation, conduct and analysis of the study.

## Supplemental Material

**Supplementary Table S1.** Demographic characteristics of study participants.

|  | **Healthy good sleepers**<br>(sublingual DEX intake) | **Healthy poor sleepers**<br>(buccal DEX intake) | **p value** |
| --- | --- | --- | --- |
| Sample size (n) | 8 | 17 |  |
| Sex ratio (male / female) | 8 / 0 | 17 / 0 |  |
| Age (years) | 23.1 ± 3.6 | 24.4 ± 3.4 | 0.408 |
| Height (cm) | 181.9 ± 9.5 | 176.9 ± 6.47 | 0.208 |
| Weight (kg) | 74.9 ± 9.7 | 70.0 ± 6.5 | 0.224 |
| Body-mass-index (kg/m<sup>2</sup>) | 22.6 ± 2.0 | 22.4 ± 1.8 | 0.778 |
| Trait anxiety | 42.1 ± 5.1 | 45.7 ± 5.8 | 0.753 |

Values indicate means ± standard deviations. Trait anxiety: score on the State-Trait Anxiety
Inventory of Spielberger <sup>1</sup>. The two groups did not differ in any of the reported variables as assessed using an independent samples t-test.

**Supplementary Table S2.** Visually scored sleep variables.

| **Sublingual ODT intake \| n=8 \| (Healthy good sleepers)** |  |  |  |
| --- | --- | --- | --- |
| **Sleep variable** | **Placebo** | **DEX<sub>20</sub> µg** | **DEX<sub>40</sub> µg** |
| Total sleep time | 430.75±17.35 | 441.69±28.59 | 444.25±13.97 |
| Sleep efficiency | 90.31±3.53 | 91.83±5.55 | 92.22±2.90 |
| Sleep latency | 13.75±10.06 | 9.68±6.25 | 10.81±6.24 |
| REM sleep latency | 92.12±16.53 | 172.19±54.79 ** | 189.88±56.87 ** |
| WASO | 33.50±11.21 | 30.13±21.10 | 28.25±13.56 |
| N1 sleep | 35.69±10.36 | 30.75±11.59 | 34.88±7.07 |
| N2 sleep | 227.50±17.10 | 245.31±26.68 (*) | 245.50±28.05 (*) |
| N3 sleep | 78.19±24.9 | 84.81±13.65 | 89.19±28.58 |
| N2 & N3 sleep | 305.69±25.25 | 330.13±29.82 * | 334.69±28.57 ** |
| REM sleep | 89.38±15.40 | 80.81±11.77 | 74.69±17.05 (*) |
| **Buccal ODT intake \| n=17 \| (Healthy poor sleepers)** |  |  |  |
| **Sleep variable** | **Placebo** | **DEX<sub>20</sub> µg** | **DEX<sub>40</sub> µg** |
| Total sleep time | 393.12 ± 42.08 | 413.27 ± 44.07 | 406.59 ± 51.40 |
| Sleep efficiency | 81.95 ± 8.88 | 86.35 ± 9.32 | 84.73 ± 10.7 |
| Sleep latency | 32.41 ± 17.99 | 21.73 ± 14.98 (*) | 21.09 ± 15.03 * |
| REM sleep latency | 125.68 ± 49.34 | 173.87 ± 45.75 ** | 242.22 ± 80.02 ***, ## |
| WASO | 54.76 ± 40.91 | 44.27 ± 44.06 | 52.47 ± 41.43 |
| N1 sleep | 30.47 ± 15.87 | 30.63 ± 18.58 | 25.59 ± 13.25 |
| N2 sleep | 204.76 ± 31.19 | 232.10 ± 33.67 * | 231.88 ± 36.34 * |
| N3 sleep | 85.15 ± 32.37 | 88.80 ± 36.27 | 95.72 ± 35.66 |
| N2 & N3 sleep | 289.91 ± 35.76 | 320.90 ± 45.66 * | 327.59 ± 37.80 * |
| REM sleep | 72.74 ± 30.16 | 61.73 ± 21.05 | 53.41 ± 28.91 (*) |

Reported values indicate means ± standard deviations in minutes, except sleep efficiency that refers to the percentage of total sleep time per time-in-bed (constant: 480 min), for the sublingual and buccal ODT formulations. Total sleep time = combined stages N1, N2, N3 and REM sleep. WASO = wakefulness after sleep onset.
\*\*\* p < 0.001, \*\* p < 0.01, \* p < 0.05, (\*) p < 0.1 (DEX vs. placebo); ## p < 0.01 (20 µg vs. 40 µg).

**Supplementary Table S3.**
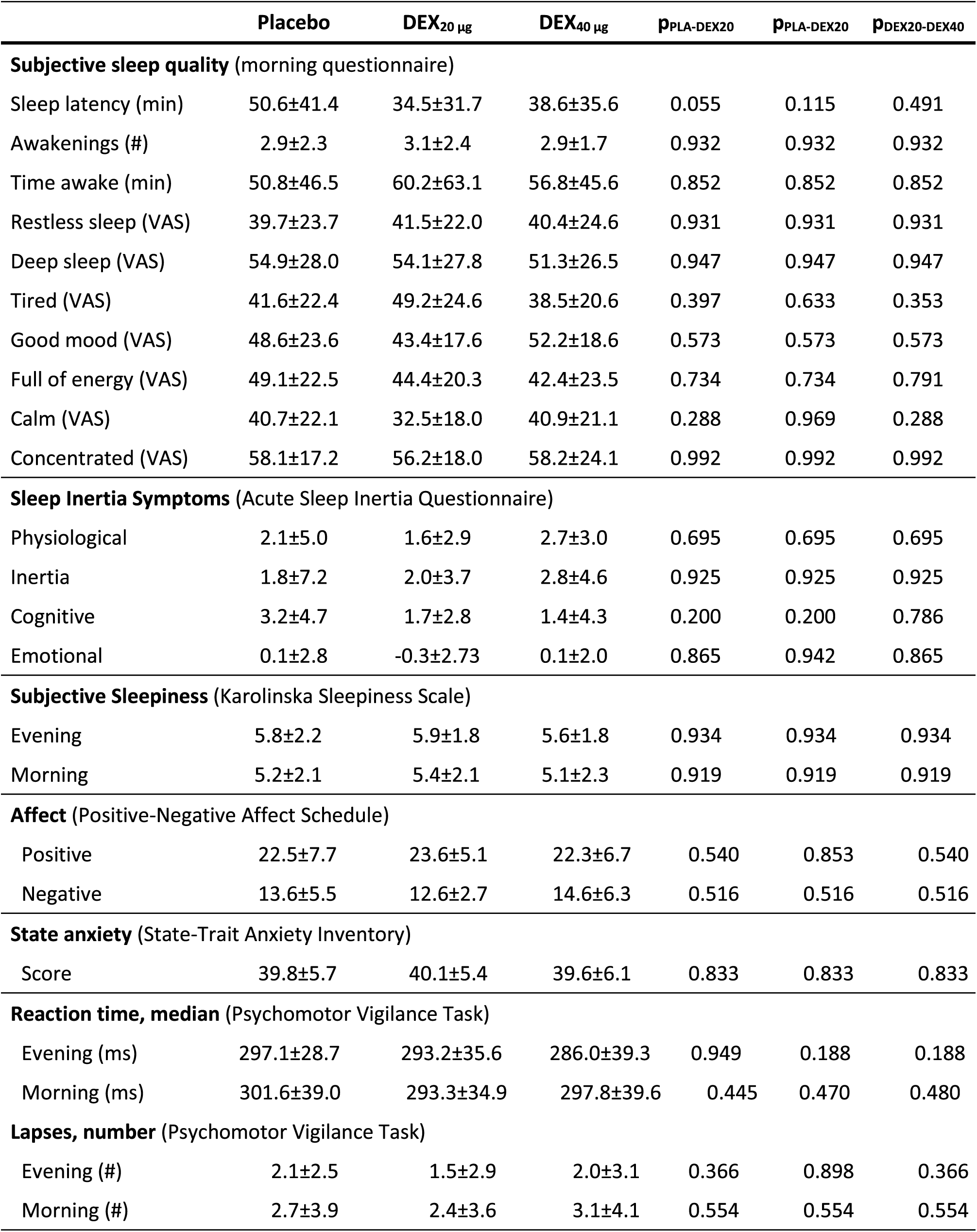
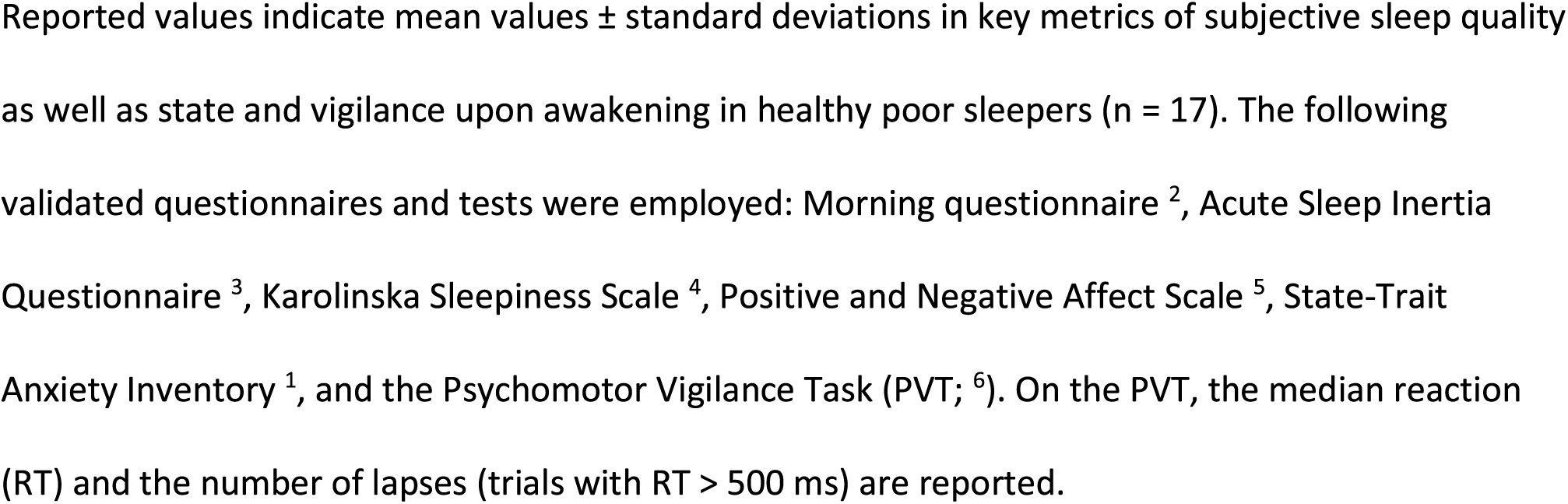
Subjective sleep quality, self-rated state and vigilance upon awakening after buccal ODT intake.

|  | Placebo | DEX<sub>20</sub> µg | DEX<sub>40</sub> µg | p<sub>PLA-DEX20</sub> | p<sub>PLA-DEX20</sub> | p<sub>DEX20-DEX40</sub> |
| --- | --- | --- | --- | --- | --- | --- |
| **Subjective sleep quality** (morning questionnaire) |  |  |  |  |  |  |
| Sleep latency (min) | 50.6±41.4 | 34.5±31.7 | 38.6±35.6 | 0.055 | 0.115 | 0.491 |
| Awakenings (#) | 2.9±2.3 | 3.1±2.4 | 2.9±1.7 | 0.932 | 0.932 | 0.932 |
| Time awake (min) | 50.8±46.5 | 60.2±63.1 | 56.8±45.6 | 0.852 | 0.852 | 0.852 |
| Restless sleep (VAS) | 39.7±23.7 | 41.5±22.0 | 40.4±24.6 | 0.931 | 0.931 | 0.931 |
| Deep sleep (VAS) | 54.9±28.0 | 54.1±27.8 | 51.3±26.5 | 0.947 | 0.947 | 0.947 |
| Tired (VAS) | 41.6±22.4 | 49.2±24.6 | 38.5±20.6 | 0.397 | 0.633 | 0.353 |
| Good mood (VAS) | 48.6±23.6 | 43.4±17.6 | 52.2±18.6 | 0.573 | 0.573 | 0.573 |
| Full of energy (VAS) | 49.1±22.5 | 44.4±20.3 | 42.4±23.5 | 0.734 | 0.734 | 0.791 |
| Calm (VAS) | 40.7±22.1 | 32.5±18.0 | 40.9±21.1 | 0.288 | 0.969 | 0.288 |
| Concentrated (VAS) | 58.1±17.2 | 56.2±18.0 | 58.2±24.1 | 0.992 | 0.992 | 0.992 |
| **Sleep Inertia Symptoms** (Acute Sleep Inertia Questionnaire) |  |  |  |  |  |  |
| Physiological | 2.1±5.0 | 1.6±2.9 | 2.7±3.0 | 0.695 | 0.695 | 0.695 |
| Inertia | 1.8±7.2 | 2.0±3.7 | 2.8±4.6 | 0.925 | 0.925 | 0.925 |
| Cognitive | 3.2±4.7 | 1.7±2.8 | 1.4±4.3 | 0.200 | 0.200 | 0.786 |
| Emotional | 0.1±2.8 | -0.3±2.73 | 0.1±2.0 | 0.865 | 0.942 | 0.865 |
| **Subjective Sleepiness** (Karolinska Sleepiness Scale) |  |  |  |  |  |  |
| Evening | 5.8±2.2 | 5.9±1.8 | 5.6±1.8 | 0.934 | 0.934 | 0.934 |
| Morning | 5.2±2.1 | 5.4±2.1 | 5.1±2.3 | 0.919 | 0.919 | 0.919 |
| **Affect** (Positive-Negative Affect Schedule) |  |  |  |  |  |  |
| Positive | 22.5±7.7 | 23.6±5.1 | 22.3±6.7 | 0.540 | 0.853 | 0.540 |
| Negative | 13.6±5.5 | 12.6±2.7 | 14.6±6.3 | 0.516 | 0.516 | 0.516 |
| **State anxiety** (State-Trait Anxiety Inventory) |  |  |  |  |  |  |
| Score | 39.8±5.7 | 40.1±5.4 | 39.6±6.1 | 0.833 | 0.833 | 0.833 |
| **Reaction time, median** (Psychomotor Vigilance Task) |  |  |  |  |  |  |
| Evening (ms) | 297.1±28.7 | 293.2±35.6 | 286.0±39.3 | 0.949 | 0.188 | 0.188 |
| Morning (ms) | 301.6±39.0 | 293.3±34.9 | 297.8±39.6 | 0.445 | 0.470 | 0.480 |
| **Lapses, number** (Psychomotor Vigilance Task) |  |  |  |  |  |  |
| Evening (#) | 2.1±2.5 | 1.5±2.9 | 2.0±3.1 | 0.366 | 0.898 | 0.366 |
| Morning (#) | 2.7±3.9 | 2.4±3.6 | 3.1±4.1 | 0.554 | 0.554 | 0.554 |

Reported values indicate mean values ± standard deviations in key metrics of subjective sleep quality as well as state and vigilance upon awakening in healthy poor sleepers ( $n = 17$ ). The following validated questionnaires and tests were employed: Morning questionnaire <sup>2</sup>, Acute Sleep Inertia Questionnaire <sup>3</sup>, Karolinska Sleepiness Scale <sup>4</sup>, Positive and Negative Affect Scale <sup>5</sup>, State-Trait Anxiety Inventory <sup>1</sup>, and the Psychomotor Vigilance Task (PVT; <sup>6</sup>). On the PVT, the median reaction (RT) and the number of lapses (trials with RT > 500 ms) are reported.

